# Substituting Blood-Based Biomarkers for Imaging Measures in Alzheimer’s Disease Studies: Implications for Sample Size and Bias

**DOI:** 10.1101/2025.11.06.25339696

**Authors:** Sarah F. Ackley, Renaud La Joie, Michelle Caunca, Shubhabrata Mukherjee, Seo-Eun Choi, Emily H. Trittschuh, Paul K. Crane, Eleanor Hayes-Larson, the Alzheimer’s Disease Neuroimaging Initiative

## Abstract

**Background:** Blood-based biomarkers for Alzheimer’s disease (AD) pathology are appealing options in large population-based studies due to their low cost, minimal invasiveness, and feasibility of collection in non-clinical settings. Despite these benefits, blood-based biomarkers have lower test-retest reliability than neuroimaging measures like amyloid positron emission tomography (amyloid-PET) Centiloids; trade-offs in power and bias remain unexplored.

**Methods:** We use data from Alzheimer’s Disease Neuroimaging Initiative (ADNI) and the Anti-Amyloid Treatment in Asymptomatic Alzheimer’s Disease (A4) studies, which include both amyloid-PET and blood-based measures, to assess differences in statistical power, required sample size, and bias when replacing a neuroimaging measure with a blood-based measure. We use simulations parameterized based on these studies to show potential implications of using plasma p-tau181 or p-tau217, blood-based AD biomarkers, in place of Centiloids from amyloid-PET, when the biomarker is either the exposure or the outcome in an analysis of interest.

**Results:** We demonstrated that substituting amyloid-PET Centiloids with a blood-based measure of p-tau can substantially reduce power, requiring 3 to 6 times the sample size to achieve 80% power compared to amyloid-PET. In addition, using a blood-based biomarker as the exposure can introduce significant regression dilution bias, attenuating estimated associations.

**Conclusions:** Due to their lower cost and ease of collection compared with neuroimaging, blood-based biomarkers facilitate AD pathology measures on larger, more diverse samples with longitudinal follow-up. Consideration of the sample sizes they necessitate and their potential for bias is critical for the design and interpretation of studies employing these biomarkers.

## Introduction

Given the growing disease burden associated with dementia and inequitable distribution across groups defined by social factors like race and education, scalable solutions for measuring disease pathology in research studies while facilitating greater inclusion and longitudinal follow-up are urgently needed (1,2). Collection of neuroimaging (e.g., positron emission tomography with amyloid ligands or amyloid-PET), cerebrospinal fluid (CSF), and neuropathological measures of Alzheimer’s disease (AD) pathology in studies has historically been challenging. Neuroimaging and CSF sample collection are expensive and burdensome, and may require visits to clinics with neuroimaging facilities (3). Neuropathology is not feasible when potential study participants are uncomfortable with brain donation, and requires substantial logistics to obtain and preserve the brain quickly following death. In addition, neuropathology cannot capture how pathology changes over the lifecourse, and distinguishing between changes that contribute to death from those associated with survival is challenging. As a result, blood-based biomarkers for AD pathology are appealing and increasingly available options in population-based studies due to their low cost, minimal invasiveness, and feasibility of collection in non-clinical settings (3,4). In addition, research using blood-based biomarkers will continue to be a priority because they are increasingly available and used for clinical practice, including diagnosis and treatment decision-making regarding amyloid-targeting drugs (5,6).

Despite the benefits of blood-based biomarkers for inclusion in longitudinal research, there are several disadvantages compared with neuroimaging measures like amyloid-PET. Neuroimaging can provide more detailed information, including topographical data about the location of pathology or atrophy in the brain. Although documentation in the literature is limited, the summary numeric measures (e.g., Centiloids, a standardized scale for brain amyloid burden designed to overcome differences across analysis techniques and tracers by anchoring scores to 0 for individuals who are confidently classified as amyloid-negative, and 100 for a typical Alzheimer’s disease patient (7)) are likely to have good reliability (7–14). In contrast, blood-based biomarkers typically only offer a single quantitative value with no topographical information, and are subject to measurement challenges, including pre-analytical factors, lower test-retest reliability, difficulty standardizing across laboratories and assays, and, because they are measured from blood, influence from peripheral factors, such as body mass index, liver function, and kidney function (15,16).

Substantial attention in the AD biomarker literature has focused on validity and measurement challenges related to sample storage and transport, assay differences, and the impact of clinical comorbidities such as body mass index and kidney function on analyte levels (15,16). However, even in the absence of these issues, random measurement error (i.e., “noise”) brings challenges, including reduced precision and potential bias (17,18). Notably, although direct head-to-head comparisons remain limited, in models that include both imaging and plasma biomarkers, PET-derived measures (including those derived from amyloid-or tau-PET) show stronger associations with cognitive outcomes than blood-based measures alone (19,20). Extant literature comparing blood-based and PET measures typically focuses on individual-level diagnosis or progression.

Among blood-based biomarkers for AD, p-tau is the most closely correlated with brain amyloid burden, possibly because it may reflect rate of tau accumulation, which is hypothesized to be driven by cumulative amyloid burden in the dominant model of AD progression) (21). As a result, measures of plasma p-tau, particularly the 217 isoform, have emerged as a key, ubiquitously used blood-based marker, with growing evidence for its use for screening and diagnosis (22,23).

Despite clear epidemiologic evidence that noise in an outcome measure reduces power, and noise in an exposure measure can induce regression dilution bias (17,18), we are not aware of evidence related to implications of measurement error in practice in AD biomarker research, such as an evaluation of the tradeoffs between the increased sample size that blood-based biomarkers facilitate with the increased precision associated with summary neuroimaging measures. For example, if a sample of 100 is needed for an amyloid-PET neuroimaging study, how many are required for similar power for a blood-based biomarker study that uses p-tau in lieu of Centiloids, and what bias could be expected if the biomarkers are an exposure variable? A clear understanding of these trade-offs is essential for study design and planning, as well as for the interpretation of secondary analyses of existing data.

To address this gap, we leverage data from Alzheimer’s Disease Neuroimaging Initiative (ADNI) and the Anti-Amyloid Treatment in Asymptomatic Alzheimer’s Disease (A4) studies, which include both amyloid PET scans and blood-based measures. We focus on amyloid-PET Centiloids and phosphorylated tau (p-tau). We use simulations parameterized based on data from ADNI and A4 to show the potential implications of measurement error for the use of blood-based biomarkers in AD research, including for research questions in which the biomarker is either the exposure or outcome.

## Methods

### Overview

Our analysis had 3 main steps. First, we used empirical data to fit models for parameters of interest, including (a) the relationship between age and Centiloids, (b) the relationship between Centiloids and p-tau, and (c) memory score as a function of age and Centiloids. Second, we used estimates from the fitted models to generate simulated study data on age, Centiloids, p-tau, and memory score, varying sample sizes of the simulated studies and the amount of random error in both the Centiloids and p-tau measures. Finally, in the simulated data, we compared the power and bias of estimated associations. Below, we describe each step in more detail.

### 1. Empirical Data and Analysis

Data used in preparation of this article were from the Alzheimer’s Disease Neuroimaging Initiative (ADNI) database and the Anti-Amyloid Treatment in Asymptomatic Alzheimer’s Disease (A4) study. All participants provided informed consent at the time of enrollment. Institutional review board (IRB) approval was obtained at each trial site.

#### Alzheimer’s Disease Neuroimaging Initiative

The ADNI was launched in 2003 as a public-private partnership, led by Principal Investigator Michael W. Weiner, MD. The original goal of ADNI was to test whether serial magnetic resonance imaging (MRI), positron emission tomography (PET), other biological markers, and clinical and neuropsychological assessment can be combined to measure the progression of mild cognitive impairment (MCI) and early Alzheimer’s disease (AD). The current goals include validating biomarkers for clinical trials, improving the generalizability of ADNI data by increasing diversity in the participant cohort, and to provide data concerning the diagnosis and progression of Alzheimer’s disease to the scientific community. For up-to-date information, see adni.loni.usc.edu. ADNI data was obtained via an application on the Laboratory of Neuro Imaging website (adni.loni.usc.edu).

We used data from ADNI participants who were cognitively normal or had mild cognitive impairment at baseline. For the blood-based biomarker, we used plasma p-tau181 measures; p-tau181 was analyzed using the Single Molecule array (Simoa) technique, using an in-house assay developed in the Clinical Neurochemistry Laboratory, University of Gothenburg, Sweden (Sept 2, 2022). We selected the earliest plasma p-tau181 for each participant, and used amyloid PET and cognitive measures closest in date to time of plasma collection. The University of California, Berkeley processed amyloid-PET data, which was used to obtain Centiloids (6mm, Nov 1, 2024). To assess cognition, we used memory scores obtained from the Center for Psychometric Analyses in Aging and Neurodegeneration (CPAAN) at the University of Washington (June 17, 2025). CPAAN is generating co-calibrated scores for memory, executive functioning, language, and visuospatial abilities using modern psychometric approaches as the Cognition Core for the Alzheimer’s Disease Sequencing Project Phenotype Harmonization Consortium (ADSP-PHC).

These scores incorporate data from multiple cognitive tests (24) and use modern psychometric approaches that result in co-calibrated and harmonized scores across datasets. Methods detailing this approach have been published (24,25).

#### A4 Study

The A4 study was a phase 3 randomized-controlled clinical trial of solanezumab among individuals with elevated amyloid assessed via amyloid-PET but without clinical symptoms. Enrollment began in 2014 and 1169 participants aged 65-86 were randomized to receive intravenous solanezumab or placebo. A4 study data were obtained from the A4/LEARN Study Data Package (A4studydata.org).

Our analysis uses data from visit 6 (time 0 during the placebo-controlled period), when plasma samples were collected, including p-tau217 measured using Lilly Clinical Diagnostics Laboratory assay. Neuroimaging data were collected at visit 2 (screening visit), up to 90 days prior to visit 6. Amyloid-PET standardized uptake value ratios (SUVrs) were converted to Centiloids using the following formula: Centiloids = 183.07*(Florbetapir SUVr) – 177.26.

Cognitive analyses focused on memory performance at visit 6 of the A4 study. Memory scores were co-calibrated and harmonized by the Center for Psychometric Analyses in Aging and Neurodegeneration with data from ADNI and several other aging cohorts (25), allowing findings related to memory to be directly compared across studies. Details of items used for score development are provided in the Supplement (Supplemental Tables S1 and S2). We only use data up to the point of randomization, so we do not include study arm in our analyses.

#### Empirical Analysis & Parameter Estimation

To parameterize simulations, we fit models separately to the ADNI and A4 data. For both studies, we fit a gamma distribution to the empirical age distribution. For the remaining relationships, we used generalized additive models for location, scale, and shape (GAMLSS). GAMLSS allows for mean modeling and also modeling variance and skewness as functions of parameters. Mean and standard deviation of Centiloids were modeled as linear functions of age using a skew-normal distribution. Mean and shape parameters for p-tau181 (ADNI) and p-tau217 (A4) were modeled as a linear function of Centiloids using a gamma distribution. The mean and variance of memory scores were modeled as functions of Centiloids and age using a normal distribution.

### 2. Simulation study

#### Data generation

We generated simulated data according to the diagram shown in **Figure 1**, using pre-fitted model parameters from ADNI or A4 (**Supplemental Tables 1 and 2**) for age, Centiloids, p-tau, and memory scores as described in the Empirical Analysis section. Age was simulated exogenously using a gamma distribution. True Centiloids, corresponding to unmeasured true amyloid burden, was simulated as a function of age using a skew-normal distribution. Measured Centiloids were simulated as a function of true Centiloids and Gaussian error. The blood-based biomarker p-tau (p-tau181 for ADNI, p-tau217 for A4) was simulated as a gamma distribution with mean (for true p-tau measure) and variance (for measured p-tau) as functions of “true Centiloids”.

**Figure 1:**
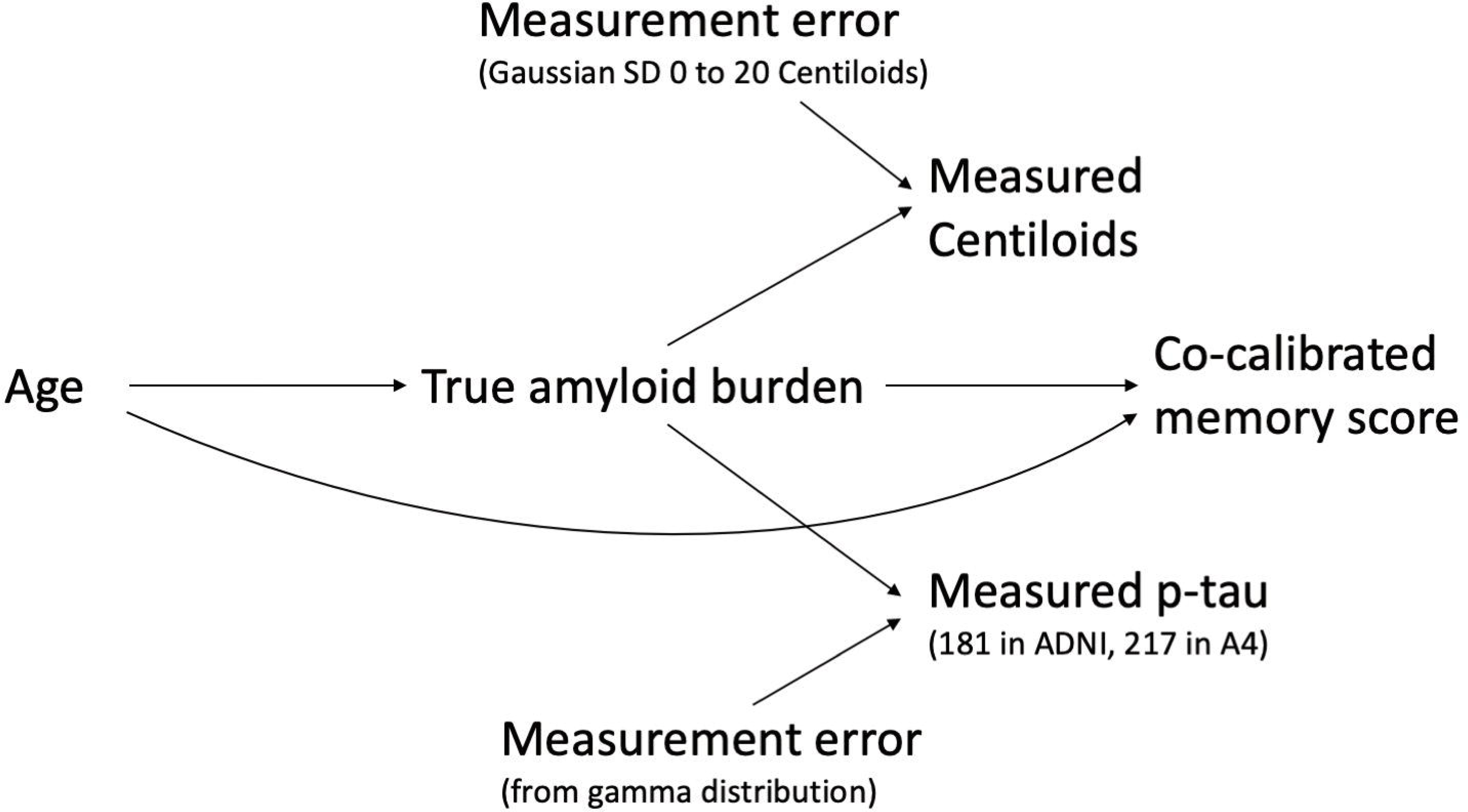
Diagram showing data-generating structure (directed acyclic graph) for simulation study parameterized based on empirical data.

Although a linear transformation of the “true” Centiloids measure was used, the “true” p-tau measure was needed to provide a comparison on the same scale as the measured p-tau to evaluate bias introduced by the variance in measured p-tau. Finally, memory scores were simulated from a normal distribution with mean and variance depending on both age and true Centiloids.

#### Scenarios

Scenarios were defined by combinations of three parameters. First, we used fitted models from either ADNI or A4 to parameterize the data generation models. Second, we varied sample sizes generated from 10 to 10,000 participants. Finally, we varied the amount of Gaussian error added to the “true” Centiloid values to obtain the “measured” Centiloid variable; for the error, we used a mean of zero and standard deviations ranging from 0 to 20 Centiloids to account for a range of plausible measurement errors. Random measurement error for Centiloids is not precisely documented in the literature. Still, it is thought to be on the order of a few Centiloids (3-12, depending on the tracer and dataset) (7,10,26,27), making our range of up to 20 Centiloids a realistic but somewhat pessimistic estimate. We evaluated a total of 110 scenarios, corresponding to parameters estimated from 2 studies times 11 sample sizes times 4 levels of noise in Centiloids and noise in p-tau.

### 3. Quantifying Power and Bias

For each scenario, we ran 5,000 data-generating iterations. In each iteration of data generation, models for the two associations of interest were estimated using standard modeling strategies. First, the relationship between age and biomarker outcome (Centiloids or p-tau) was estimated using simple linear regression. Second, the relationship between the biomarker and the memory score was estimated using linear regression adjusted for age.

Simulation results were summarized across the 5,000 iterations by scenario (i.e., study used to parameterize the simulations, sample size, and magnitude of added error). Power was calculated as the proportion of times p < 0.05 was achieved for the coefficient of interest (age, in age-biomarker models, and biomarker, in biomarker-memory score models). Confidence intervals were calculated using the exact binomial method. Bias was calculated for each iteration as the difference between a coefficient for the measured biomarker and the coefficient for the true biomarker (i.e., coefficients for Centiloids with Gaussian error added vs. no error added, or coefficients for p-tau with variance added vs. p-tau that was a linear transformation of true Centiloids). Confidence intervals were calculated by multiplying the standard norm deviate (approximately 1.96) by the standard error, where the standard error is the standard deviation of the difference divided by the square root of 5000, the number of simulations.

## Results

Descriptive statistics for the ADNI and A4 datasets are presented in **Table 1. Supplemental Figure S1** shows the distribution of Centiloid values for both cohorts. The analytic samples included 952 individuals from ADNI and 1082 individuals from A4. Mean age and co-calibrated and harmonized memory scores were comparable across the two cohorts, although memory scores were slightly higher in ADNI. Centiloids levels were higher in A4, reflecting that amyloid-PET positivity was an inclusion criterion for that study. Levels of p-tau are not directly comparable since platforms and assays differed: p-tau181 was used in ADNI, and p-tau217 was used in A4. **Supplemental Tables 3** and **4** provide the parameters for models fit to these data and used for subsequent simulations.

**Table 1:**
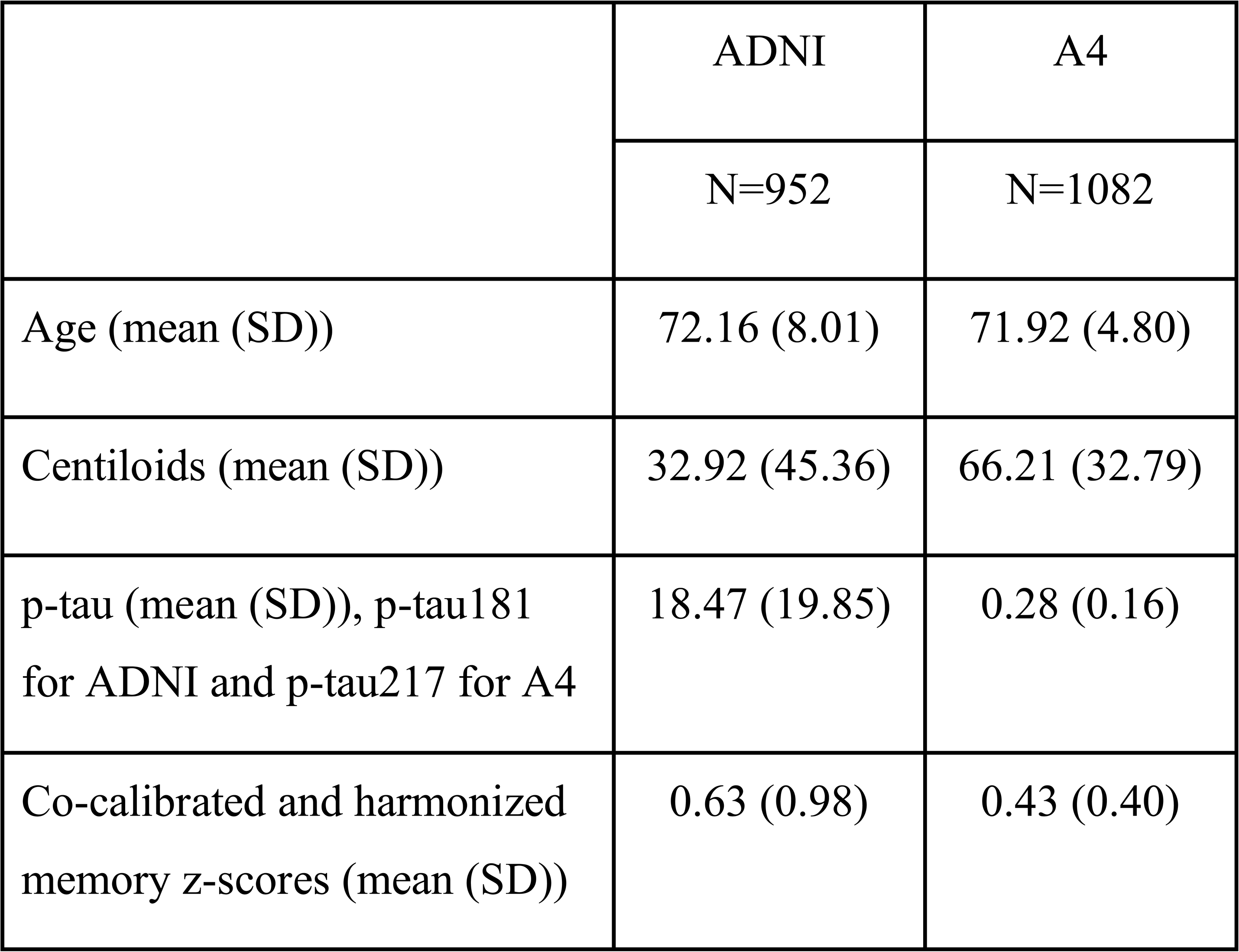
Descriptives of variables used to parameterize simulations in the ADNI and A4 analytic samples.

|  | ADNI | A4 |
| --- | --- | --- |
|  | N=952 | N=1082 |
| Age (mean (SD)) | 72.16 (8.01) | 71.92 (4.80) |
| Centiloids (mean (SD)) | 32.92 (45.36) | 66.21 (32.79) |
| p-tau (mean (SD)), p-tau181 for ADNI and p-tau217 for A4 | 18.47 (19.85) | 0.28 (0.16) |
| Co-calibrated and harmonized memory z-scores (mean (SD)) | 0.63 (0.98) | 0.43 (0.40) |

In simulations, we first evaluated bias and power for estimated associations between age (independent variable) and the biomarker (dependent variable). As shown in **Figure 2A**, additional measurement error in Centiloids up to 20 Centiloids did not have a large impact on power in either cohort. However, as also shown in **Figure 2A**, substituting plasma p-tau for Centiloids significantly reduced statistical power. **Figure 2B** shows that the difference in power was larger in ADNI than in A4. **Figure 2C** shows that sample sizes required for 80% power when using plasma p-tau were larger in ADNI than in A4. Achieving 80% power required 3 to 6 times the sample size when using plasma p-tau measures versus the true Centiloids measure (Tabular data in **Supplemental Table S3**). **Supplemental Figure S2** shows that there was no bias in the estimated coefficient for age in models with a biomarker outcome.

**Figure 2:**
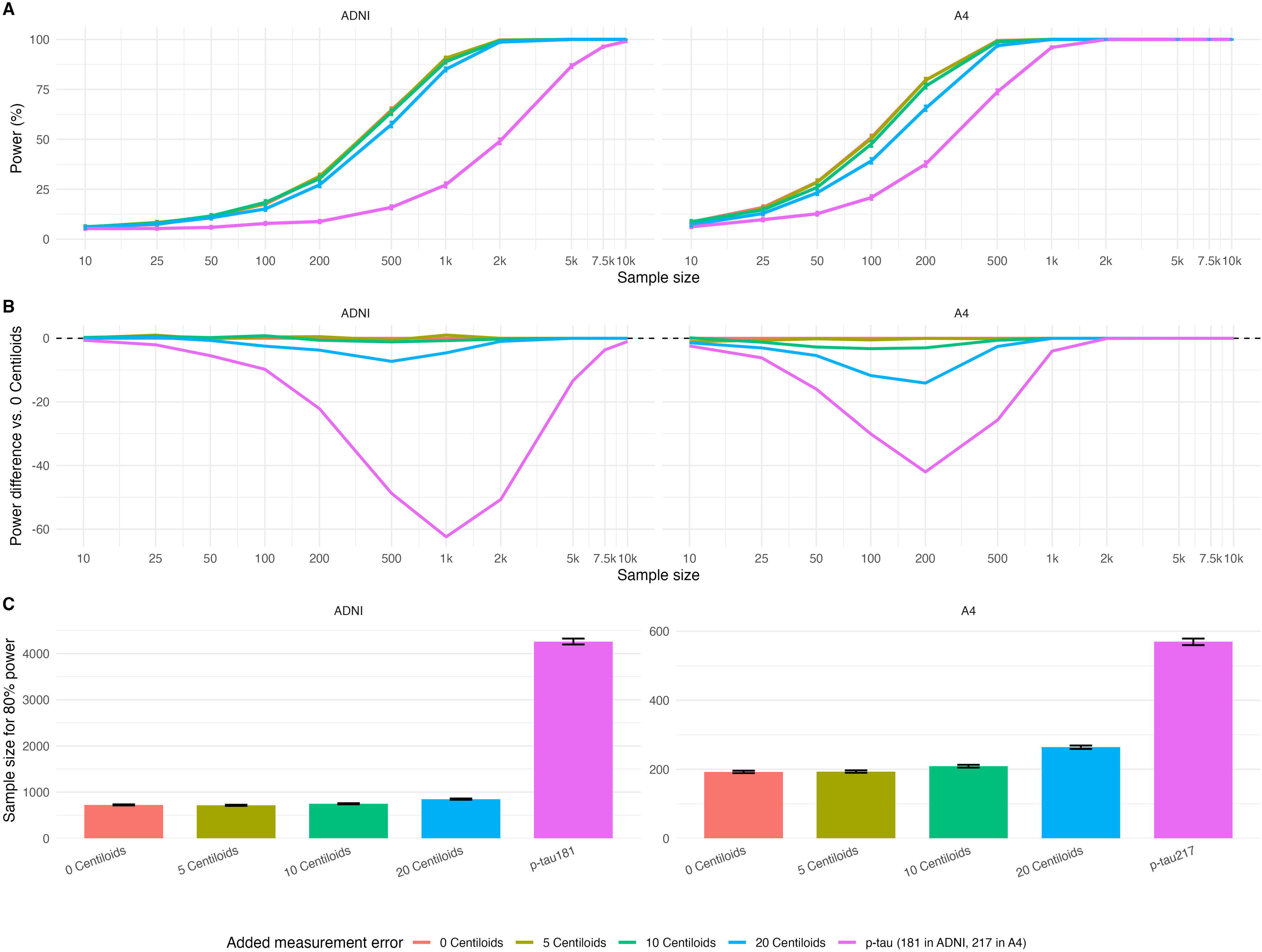
Power curves and power curve differences for biomarker outcome models. **A.** Percent power as a function of sample size for detecting a statistically significant correlation between a biomarker and age. Power curves for ADNI and A4 show that power is not hugely affected by added error in Centiloids, but replacing Centiloids with p-tau concentrations dramatically reduces power. **B.** Power curve differences show the difference in power compared with using true Centiloids (no added error) as the outcome measure. Reductions in power when replacing Centiloids with p-tau is larger in ADNI than in A4. **C.** Number of participants needed to achieve 80% power for true Centiloids, Centiloids with added error, and p-tau in ADNI and A4 in biomarker outcome models. For models assessing the relationship between an exposure, age, and a biomarker, the sample size required to achieve 80% power is between 3 and 6 times larger when p-tau vs. Centiloids is the outcome. 95% confidence intervals are calculated using the delta method.

Simulation findings differed when estimating associations between biomarkers (independent variable) and memory score (dependent variable), adjusted for age. As shown in **Figure 3**, regression dilution bias attenuated estimated associations whenever measurement error was present, and increased with the magnitude of measurement error in Centiloids (e.g., for 20 Centiloids of error, bias was −17% in ADNI and −28% in A4). Negative bias was particularly substantial when estimating associations using plasma p-tau (−87% for p-tau181 in ADNI and - 62% for p-tau217 in A4). As shown in **Figure 4**, measurement error was also associated with substantial decreases in power to detect statistically significant biomarker coefficients, a consequence of the attenuation of the estimated association due to regression dilution bias.

**Figure 3:**
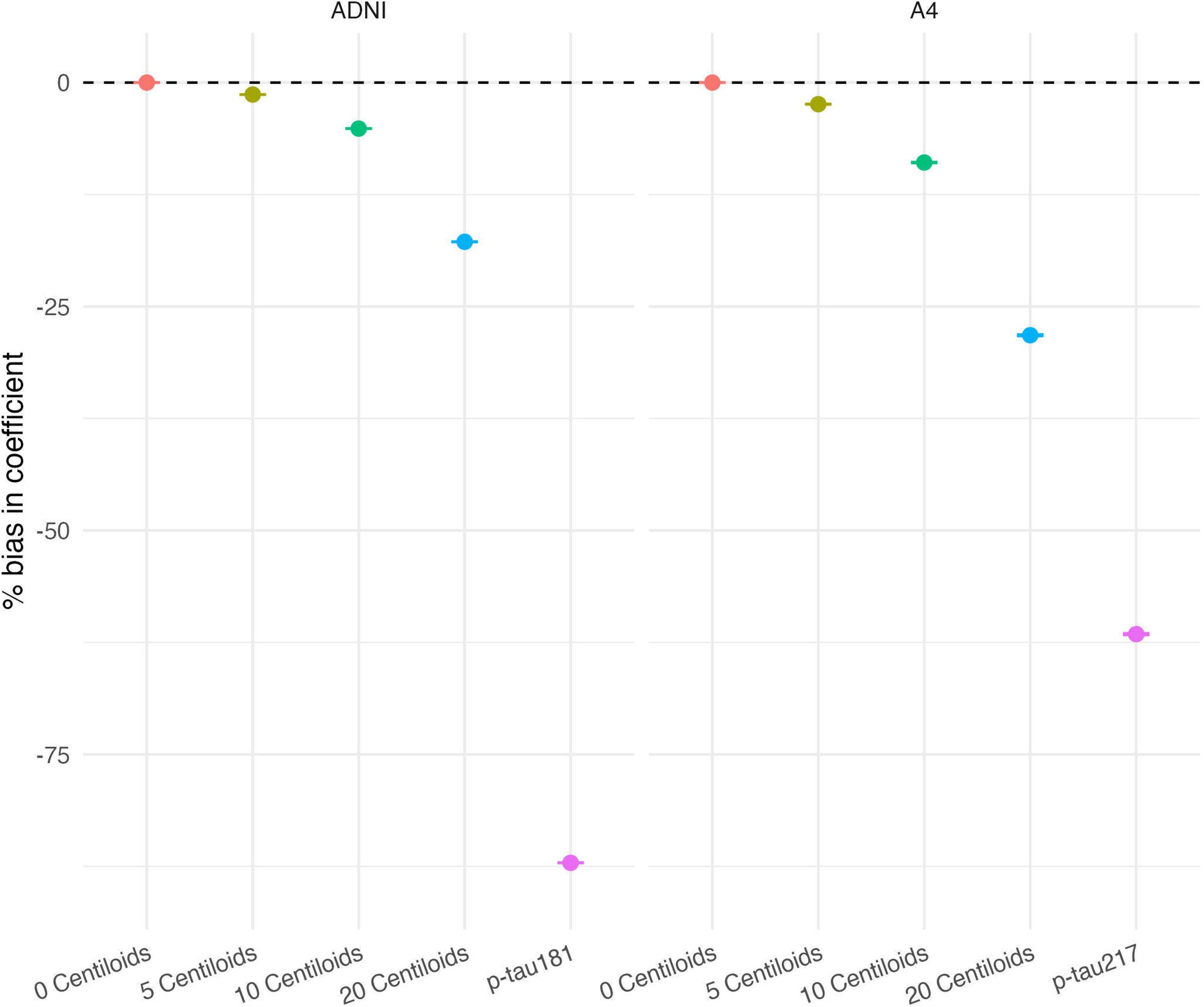
Bias under each scenario for models with the biomarker-as-exposure and memory score as the outcome for ADNI and A4. Bias is estimated using simulations with sample sizes of 10,000. Adding error to true Centiloids only modestly attenuates association estimates, whereas replacing Centiloids with p-tau dramatically attenuates association estimates in both cohorts.

**Figure 4:**
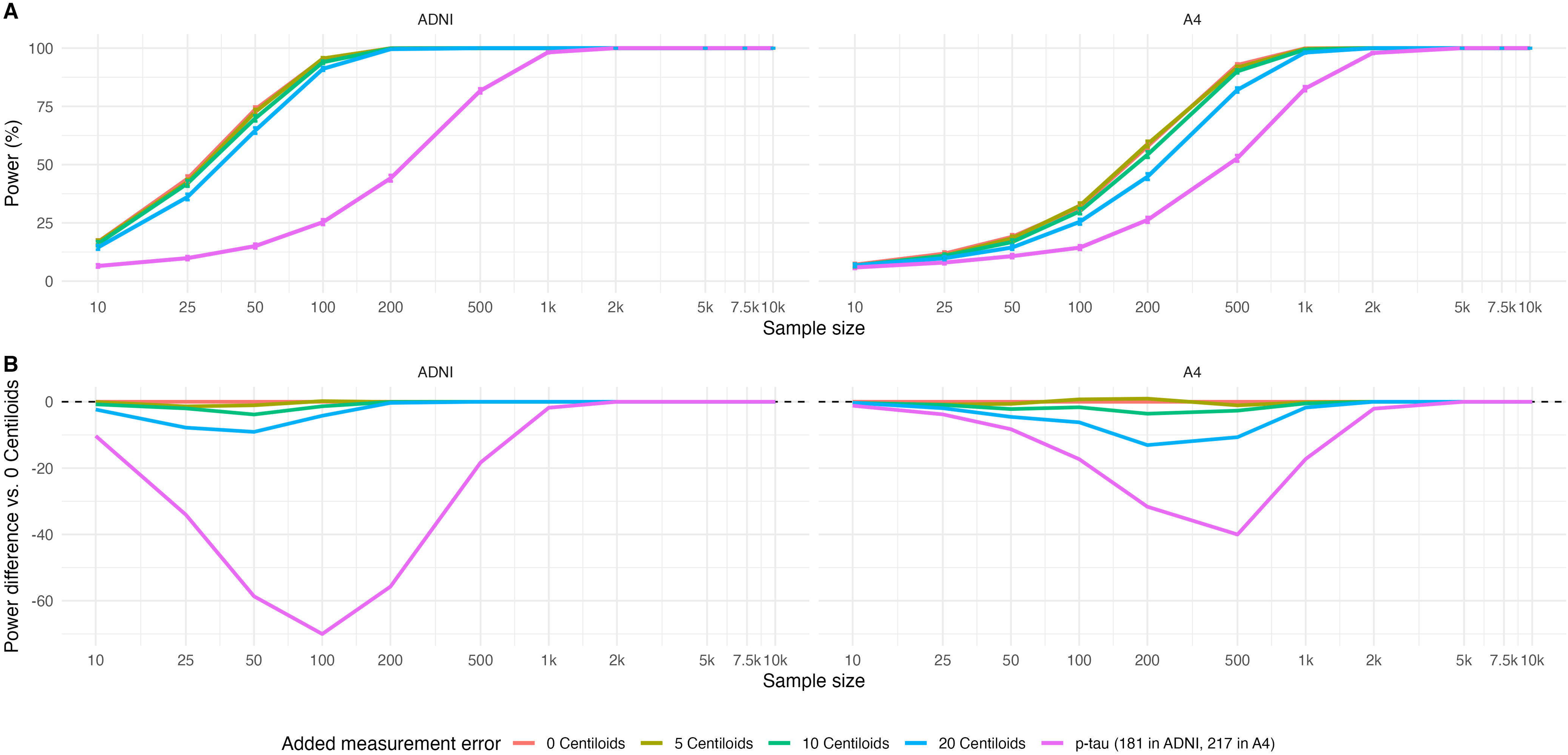
Power curves for biomarker-as-exposure models. In addition to inducing regression dilution bias, measurement error reduces power in biomarker-as-exposure models. Dramatic reductions in power are seen when using plasma p-tau, with larger reductions in ADNI. **A.** Percent power as a function of sample size for detecting a statistically significant coefficient for the biomarker and memory score, adjusting for age. **B.** Power curve differences show the difference in power compared with using true Centiloids (no added error) as the exposure measure. Reductions in power when replacing Centiloids with plasma p-tau is larger in ADNI than in A4.

## Discussion

The study aimed to quantify potential implications for power and bias of using blood-based AD biomarkers in place of neuroimaging measures in AD biomarker studies. We demonstrated that using blood-based p-tau in lieu of amyloid-PET Centiloids can substantially reduce power to detect associations when the biomarker is the outcome, and can introduce significant regression dilution bias, yielding attenuated association estimates and reduced power when the biomarker is the exposure. Relationships between age and amyloid and amyloid and memory score depended significantly on the cohort, but p-tau217 (Lilly) in the A4 study appeared to outperform p-tau181 (in-house Simoa) in the ADNI relative to Centiloids.

There is significant and warranted enthusiasm surrounding AD blood-based biomarkers in both research and clinical practice. Epidemiologists have long called for more representative samples, including in AD research (28); until recently (29), the majority of neuroimaging cohorts have comprised predominantly non-Latino White populations. Blood-based biomarkers have significant appeal for feasibly increasing the diversity and representativeness of samples and for longitudinal follow-up, as they are less expensive and do not require travel to a neuroimaging site, typically an academic medical center (3). There are additional geographic requirements for proximity to equipment for generating radioactive tracers used in PET imaging, making PET infeasible in many rural settings (30–32). In contrast, blood can be collected at a wider range of locations, including in-home collection or, as the field develops further, potentially as dried blood spots (33); samples can then be shipped to and processed at a central laboratory (34).

Clinically, recommendations already exist for using blood-based biomarkers for diagnosis and to make prognoses about dementia risk (35,36). Following the approvals of the amyloid-targeting therapies lecanemab and donanemab, this enthusiasm has grown, as these biomarkers currently play and will continue to play a role in determining eligibility for treatment (6). Approaches to account for measurement error and uncertainty in clinical use are in development (e.g. a two-threshold approach with very low values ruling out amyloid positivity, very high values ruling it in, and follow-up imaging for those with intermediate values) (37); ongoing research and integration with other biomarkers and diagnostics is going to remain a significant priority.

In the research setting, our results show that studies using p-tau measures need to be substantially larger to achieve the same statistical power as a comparable amyloid-PET study (3 to 6 times larger for the relationships we examined in this analysis, based on ADNI and A4). Even allowing for pessimistic measurement error in Centiloids (i.e., 20 Centiloids of error), sample sizes required for comparable 80% power analyses using plasma p-tau were 2 to 5 times larger.

Absolute power to detect age-amyloid relationships was lower, and differences between power using p-tau versus centiloids measures were greater in ADNI than A4, likely reflecting both the difference between p-tau181 (ADNI) and p-tau217 (A4) and the weaker age-Centiloid relationship in the empirical data in ADNI than in A4.

We also observed substantial reductions in power and large bias (attenuation) up to 85% for measures of p-tau versus Centiloids when the biomarker was the exposure of interest. Exact magnitudes of bias and reduced power in other settings likely depend on the sample composition, associations, and biomarkers of interest. Still, it is possible that both reduced power and regression dilution bias may explain the mixed or null findings in several recent AD biomarker studies (38,39). As a result, researchers using existing biomarker data should consider quantitative bias analysis to evaluate the potential role of measurement error in their findings (40,41). In addition, using combinations of multiple biomarkers (e.g., a factor score with better measurement precision) may reduce some of these concerns, and research on how best to integrate multiple measures is ongoing (42–44).

Our findings also have several implications for planning future research using blood-based AD biomarkers. Investigators should consider tradeoffs between the ease and accessibility of blood-based biomarkers and the larger sample size required for adequate statistical power compared with neuroimaging. Given that a panel of AD blood tests currently costs a few hundred dollars, while the cost of an amyloid-PET scan can exceed several thousand dollars, cost-effectiveness may still favor blood-based biomarkers (45). However, recruitment and evaluation of additional study participants confer additional costs, particularly in longitudinal studies. In addition, increasing sample size does not mitigate regression dilution bias, and researchers may want to consider the collection of amyloid-PET imaging in a subset to develop quantitative methods to account for such bias. Careful consideration of such issues in the study design phase can ensure optimal use of study resources (46).

This study has several limitations. We used a simulation approach, which has the key advantage of allowing specification and comparison against a known “truth,” but requires simplifications to isolate the question of interest, in this case related to measurement error in biomarkers. For example, our approach assumed that Centiloids was an unbiased gold standard. To mitigate this, we included scenarios in which Centiloids are also measured with random error up to 20 Centiloids, which is larger (more pessimistic) than suggested in the literature (7–14); other forms of systematic error, including atrophy and partial volume effects (47,48), were beyond the scope of this paper. In addition, we treated errors in Centiloids and variability in plasma p-tau relative to Centiloids as random and homoskedastic measurement errors. Non-random measurement error may reasonably be modeled as random, such as error due to small differences in thaw time across samples that do not systematically differ by study site or participant characteristics.

Systematic measurement error in either Centiloids or p-tau that is not reasonably modeled as random, which we do not address, may induce additional biases. We did not account for other factors that may be associated with p-tau levels such as BMI, liver function, or kidney function, either through adjustment or use of a p-tau to total tau ratio (15,49); although these may account for some variability in p-tau relative to Centiloids, making our estimates of measurement error in p-tau potentially somewhat pessimistic, the field is still developing best practices and standardized approaches.

We also did not account for measurement error in other variables (e.g., memory score); however, the co-calibrated memory score we used has been shown to have lower measurement error than single cognitive tests (50). We expect that absolute power would be lower in all scenarios for cognitive outcomes with lower measurement precision. For example, the harmonized memory score has lower precision in A4 than in ADNI, and this is likely a driver of the lower power at a given sample size in Figure 5 for A4 than ADNI. Implications for differences in power between Centiloids and p-tau measures based on cognitive outcome measurement precision are harder to predict. Finally, we focused on isolating the impact of measurement error on estimated associations, rather than causal effects, and therefore included minimal confounder adjustment beyond age and did not evaluate potential selection biases.

In addition, our analysis was limited to demonstrating the potential impact of measurement error in blood-based biomarkers using parameters derived from specific empirical data; although the principles underlying the measurement error apply broadly, our specific findings may not generalize to other cohorts or measures. This is clearly demonstrated by differences in our results between the ADNI and A4 studies; these studies have different inclusion criteria, different Centiloids distributions, use different plasma p-tau measures (p-tau181 in ADNI, p-tau217 in A4), have varying strengths of association for variables of interest, and consequently show different sample size requirements and magnitude of bias. In addition, we considered only plasma p-tau, which is generally regarded as outperforming other biomarkers due to its higher specificity compared with GFAP and NfL, as well as its improved performance relative to Aβ42/40 (the ratio of amyloid beta 42 to amyloid beta 40) in predicting amyloid Centiloids, but measurement error and its impact likely differ across different biomarkers, or across different assays for a given biomarker. Researchers should consider the specific properties of a given measure when evaluating the generalizability of our findings to their work.

In summary, despite their expanding role in clinical practice and great promise for capturing AD pathology in large population-based research samples, we showed that plausible magnitudes of measurement error in blood-based biomarkers could substantially reduce power and introduce regression dilution bias, attenuating estimated effects. Awareness of these challenges can facilitate optimal design, analysis, and interpretation of studies employing these biomarkers.

## Data Availability

For up-to-date information, see adni.loni.usc.edu. ADNI data was obtained via an application on the Laboratory of Neuro Imaging website (adni.loni.usc.edu).
A4 study data were obtained from the A4/LEARN Study Data Package (A4studydata.org).

https://adni.loni.usc.edu/

https://www.a4studydata.org/

## Acknowledgments

Data collection and sharing for this project was funded by the Alzheimer’s Disease Neuroimaging Initiative (ADNI) (National Institutes of Health Grant U01 AG024904) and DOD ADNI (Department of Defense award number W81XWH-12-2-0012). ADNI is funded by the National Institute on Aging, the National Institute of Biomedical Imaging and Bioengineering, and through generous contributions from the following: AbbVie, Alzheimer’s Association; Alzheimer’s Drug Discovery Foundation; Araclon Biotech; BioClinica, Inc.; Biogen; Bristol-Myers Squibb Company; CereSpir, Inc.; Cogstate; Eisai Inc.; Elan Pharmaceuticals, Inc.; Eli Lilly and Company; EuroImmun; F. Hoffmann-La Roche Ltd and its affiliated company Genentech, Inc.; Fujirebio; GE Healthcare; IXICO Ltd.; Janssen Alzheimer Immunotherapy Research & Development, LLC.; Johnson & Johnson Pharmaceutical Research & Development LLC.; Lumosity; Lundbeck; Merck & Co., Inc.; Meso Scale Diagnostics, LLC.; NeuroRx Research; Neurotrack Technologies; Novartis Pharmaceuticals Corporation; Pfizer Inc.; Piramal Imaging; Servier; Takeda Pharmaceutical Company; and Transition Therapeutics. The Canadian Institutes of Health Research is providing funds to support ADNI clinical sites in Canada. Private sector contributions are facilitated by the Foundation for the National Institutes of Health (www.fnih.org). The grantee organization is the Northern California Institute for Research and Education, and the study is coordinated by the Alzheimer’s Therapeutic Research Institute at the University of Southern California. ADNI data are disseminated by the Laboratory for Neuro Imaging at the University of Southern California.

The A4 Study was a secondary prevention trial in preclinical Alzheimer’s disease, aiming to slow cognitive decline associated with brain amyloid accumulation in clinically normal older individuals. The A4 Study was funded by a public-private-philanthropic partnership, including funding from the National Institutes of Health-National Institute on Aging, Eli Lilly and Company, Alzheimer’s Association, Accelerating Medicines Partnership, GHR Foundation, an anonymous foundation, and additional private donors, with in-kind support from Avid Radiopharmaceuticals, Cogstate, Albert Einstein College of Medicine and the Foundation for Neurologic Diseases.The companion observational Longitudinal Evaluation of Amyloid Risk and Neurodegeneration (LEARN) Study was funded by the Alzheimer’s Association and GHR Foundation. The A4 and LEARN Studies were led by Dr. Reisa Sperling at Brigham and Women’s Hospital, Harvard Medical School, and Dr. Paul Aisen at the Alzheimer’s Therapeutic Research Institute (ATRI) at the University of Southern California. The A4 and LEARN Studies were coordinated by ATRI at the University of Southern California, and the data are made available under the auspices of Alzheimer’s Clinical Trial Consortium through the Global Research & Imaging Platform (GRIP). The complete A4 Study Team list is available on: https://www.actcinfo.org/a4-study-team-lists/. We would like to acknowledge the dedication of the study participants and their study partners who made the A4 and LEARN Studies possible.

Calibration of memory scores for ADNI and for A4 were funded by U24 AG074855 from NIA.

