## Supplemental Information for "Substituting Blood-Based Biomarkers for Imaging Measures in Alzheimer’s Disease Studies: Implications for Sample Size and Bias"

*Details on the Harmonized A4 Memory Score Development*

There are three steps to produce harmonized scores; (1) pre-calibration: we figure out the best model which is the most appropriate to the cross-sectional data. In other words, we try to understand the data dimensionality. The cross-sectional data consists of last available visits with age at least 60, from each participant; (2) co-calibration: if there is at least one non-anchor, which is the item without loading and thresholds estimated from previous study, we run co-calibration to estimate those parameters; (3) scoring: using all loadings and thresholds, we run the model suggested from the pre-calibration step.

There are three versions (SC, A, B) of test items across visits in A4. Here are more details.

**Pre-calibration:** We tested several models using confirmatory factor analysis (CFA) for the memory domain using the last available visit from each individual. We found that the best model was a bifactor one with four secondary structures: a) a residual correlation between each pair of story immediate recall and delayed recall Logical Memory (LM) items; and b) a residual correlation between the two MMSE items, *date* and *month*. Other items included were three versions of the Free and Cued Selective Reminding Test (FCSRT) and items from the Mini-Mental State Examination (MMSE), which included three different wordlists in addition to version-free items like *date*, *month*, *year*, etc.. A complete list of items from A4 used to derive the memory scores can be found in Table S1.

**Co-calibration:** We used the same sample for this step as laid out under pre-calibration. A CFA model with Maximum Likelihood-Robust (MLR) estimation was used to estimate unique items (non-anchors) in A4, while anchor items were set to their already estimated values in the Center for Psychometric Analyses in Aging and Neurodegeneration (CPAAN) item bank, which includes item parameters from aging studies, including ADNI. A list of anchor and non-anchor items can be found in Table S2.

**Scoring:** We used a CFA model with item parameters (loadings and thresholds) for A4 unique items from the co-calibration step and anchor item parameters from the CPAAN item bank to compute memory composite scores across all visits.

**Table S1.** A4 cognitive items used to generate harmonized memory scores. Secondary structure labels follow those from CPAAN item bank.

| Raw item | Used item | Description | Anchor | Secondary structure |
|---|---|---|---|---|
| limmtotal, ldeltotal | rlimmtot | LM immediate ver SC: Anna Thompson | yes | f2 |
| | rldeltot | LM delayed: ver SC: Anna Thompson | yes | f2 |
| | rwlm_imb | LM immediate ver SC: Robert Miller | yes | f3 |
| | rwlm_deb | LM delayed ver SC: Robert Miller | yes | f3 |
| | rlmiagf | LM immediate ver A: Greg Fortune | | f38 |
| | rlmdagf | LM delayed ver A: Greg Fortune | | f38 |
| | rlmibmj | LM immediate ver B: Martha Jackson | | f39 |
| | rlmdbmj | LM delayed ver B: Martha Jackson | | f39 |
| fcfreet1 | rfcfr1sc | FCSRT free recall ver SC: tulip | | |
| | rfcfr1a | FCSRT free recall ver A: spider | | |
| | rfcfr1b | FCSRT free recall ver B: onion | | |
| immediate: mmball, mmflag, mmtree delayed: mmballdl, mmflagdl, mmtreedl | wordimsc | MMSE ver SC (pony, quarter, orange) | | |
| | worddesc | MMSE ver SC | | |
| | ratb1 | MMSE ver A (apple, table, penny) | yes | |
| | ratb2 | MMSE ver A | yes | |
| | rbft1 | MMSE ver B (ball, flag, tree) | yes | |
| | rbft2 | MMSE ver B | yes | |
| mmdate | rmmdate | MMSE date | yes | f7 |
| mmmonth | rmmmonth | MMSE month | yes | f7 |
| mmyear | rmmyear | MMSE year | yes | |
| mmday | mmday | MMSE day of the week | yes | |
| mmseason | mmseason | MMSE season | yes | |
| mmhospit | mmhospit | MMSE mmhospit | yes | |
| mmfloor | mmfloor | MMSE floor | yes | |
| mmcity | rmmctst | MMSE mmcity + mmstate | yes | |
| mmstate | rmmctst | MMSE mmcity + mmstate | yes | |
| mmarea | mmarea | MMSE county | yes | |

**Table S2:** Anchors / non-anchors at each version

<table>
<tr><th></th><th colspan="2">version SC</th><th>version A</th><th>version B</th></tr>
<tr><td>LM</td><td>anchor (AT)</td><td>anchor (RM)</td><td>non-anchor</td><td>non-anchor</td></tr>
<tr><td>FCSRT</td><td colspan="2">non-anchor</td><td>non-anchor</td><td>non-anchor</td></tr>
<tr><td>MMSE word recall</td><td colspan="2">non-anchor</td><td>anchor</td><td>anchor</td></tr>
<tr><td>MMSE other items</td><td colspan="4">anchor</td></tr>
</table>

**Table S3.** *Estimated parameters from models used to parameterize simulations of age, amyloid (Centiloid), p-tau, and memory score based on ADNI. These fitted parameters were used to generate synthetic data for simulations.*

| **Outcome** | **Term** | **Rate** | **Shape** | **Mu** | **Sigma** | **Nu** |
|---|---|---|---|---|---|---|
| age | rate | 1.113 | | | | |
| age | shape | | 80.291 | | | |
| Centiloids | (Intercept) | | | 4.279 | 3.081 | 7.676 |
| Centiloids | age | | | -0.35 | 0.016 | 0.057 |
| p-tau | (Intercept) | | | 2.727 | -0.488 | |
| p-tau | Centiloids | | | 0.005 | | |
| memory | (Intercept) | | | 2.412 | -0.735 | |
| memory | Centiloids | | | -0.008 | 0.001 | |
| memory | age | | | -0.021 | 0.008 | |

**Table S4.** *Estimated parameters from models used to parameterize simulations of age, amyloid (Centiloid), p-tau, and memory score based on A4. These fitted parameters were used to generate synthetic data for simulations.*

| **Outcome** | **Term** | **Rate** | **Shape** | **Mu** | **Sigma** | **Nu** |
|---|---|---|---|---|---|---|
| age | rate | 3.199 | | | | |
| age | shape | | 230.107 | | | |
| Centiloids | (Intercept) | | | -42.308 | 3.158 | 8.5 |
| Centiloids | age | | | 0.965 | 0.011 | -0.058 |
| p-tau | (Intercept) | | | -1.907 | -0.942 | |
| p-tau | Centiloids | | | 0.009 | | |
| memory | (Intercept) | | | 1.771 | -1.04 | |
| memory | Centiloids | | | -0.002 | 0.001 | |
| memory | age | | | -0.017 | 0 | |

**Table S5.** Sample size (95% confidence interval) needed to achieve 80% power under varying measurement error scenarios

| Study | Noise added | N needed for 80% power | Lower CI | Upper CI |
|---|---|---|---|---|
| ADNI | 0 Centiloids | 723 | 712 | 735 |
| ADNI | 5 Centiloids | 714 | 702 | 726 |
| ADNI | 10 Centiloids | 748 | 736 | 760 |
| ADNI | 20 Centiloids | 849 | 835 | 862 |
| ADNI | p-tau181 | 4261 | 4197 | 4325 |
| A4 | 0 Centiloids | 192 | 189 | 196 |
| A4 | 5 Centiloids | 193 | 190 | 197 |
| A4 | 10 Centiloids | 209 | 205 | 213 |
| A4 | 20 Centiloids | 264 | 259 | 269 |
| A4 | p-tau217 | 570 | 560 | 579 |

**Figure S1**: Distribution of Centiloid values by cohort.

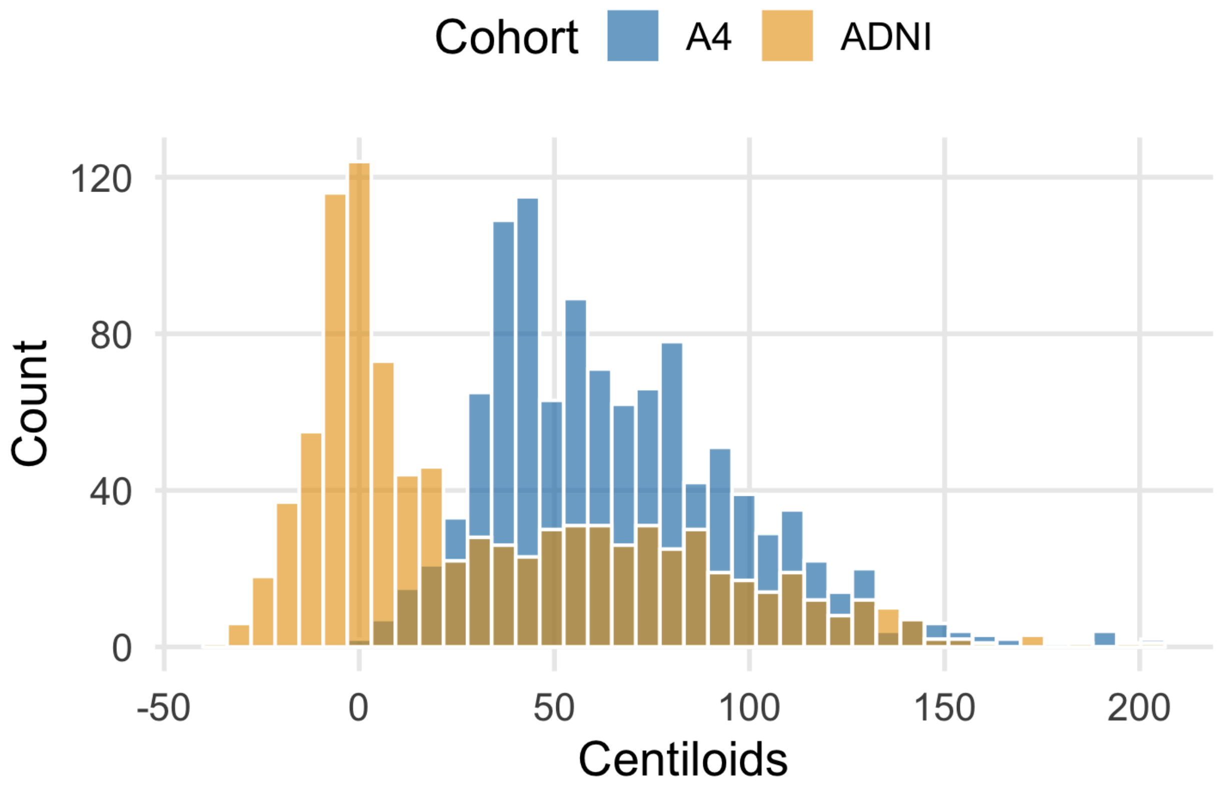

**Figure S2:** Percent bias in coefficients with a biomarker outcome. There is no bias in the estimated coefficient for age in models with a biomarker outcome.

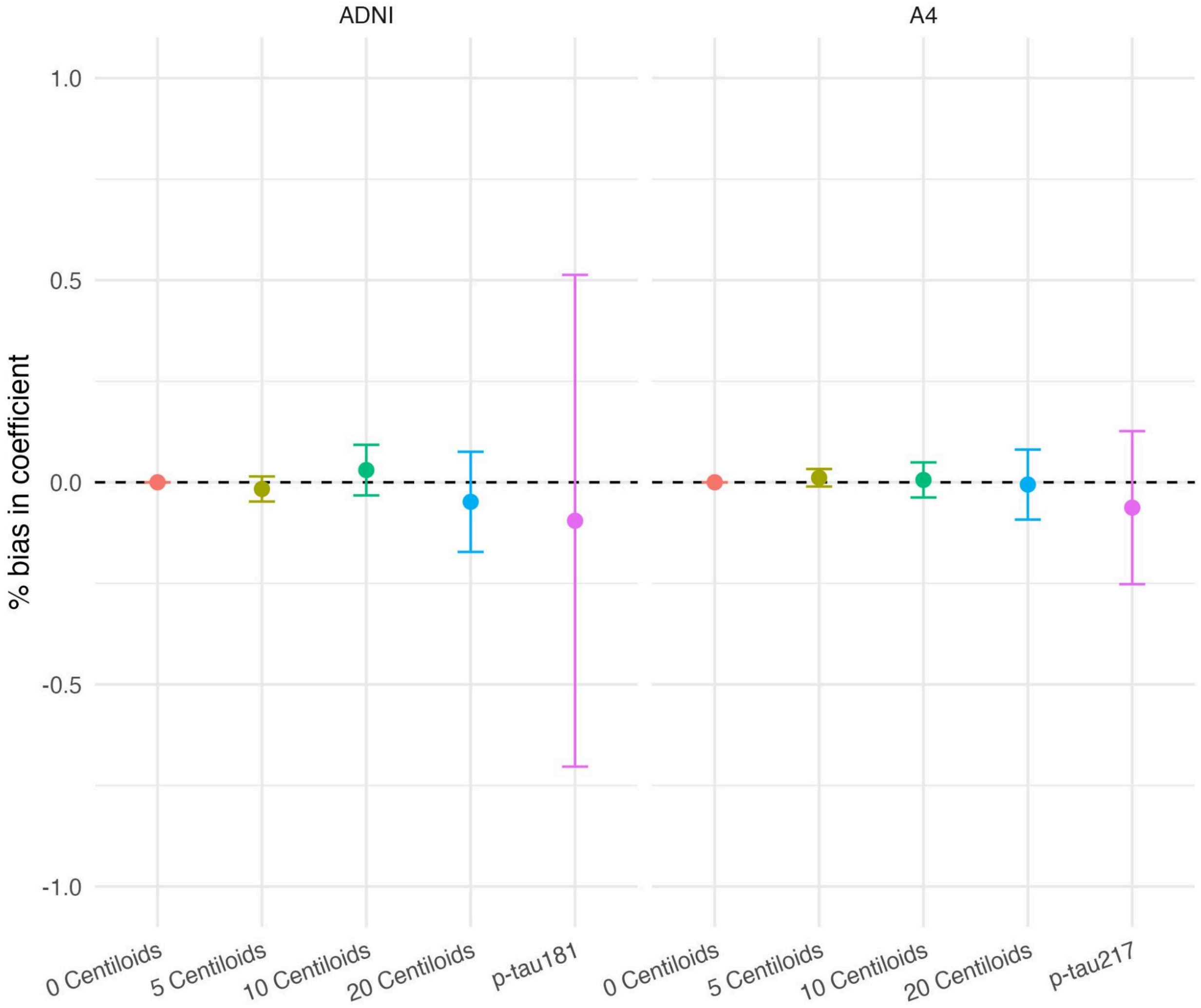
